# Lack of lockdown, open borders, and no vaccination in sight: is Bosnia and Herzegovina a control group?

**DOI:** 10.1101/2021.03.01.21252700

**Authors:** Adnan Fojnica, Ahmed Osmanovic, Nermin Đuzic, Armin Fejzic, Ensar Mekic, Zehra Gromilic, Imer Muhovic, Amina Kurtovic-Kozaric

## Abstract

Bosnia and Herzegovina is among ten countries in the world with the highest mortality rate due to COVID-19 infection. Lack of lockdown, open borders, high mortality rate, no herd immunity, no vaccination plan, and strong domestic anti-vaccination movement present serious COVID-19 concerns in Bosnia and Herzegovina. In such circumstances, we set out to study if the population is willing to receive the vaccine.

A cross-sectional study was conducted among 10,471 adults in Bosnia and Herzegovina to assess the attitude of participants toward COVID-19 vaccination. Using a logistic regression model, we assessed the associations of sociodemographic characteristics with vaccine rejection, reasons for vaccine hesitancy, preferred vaccine manufacturer, and information sources.

Surprisingly, only 25.7% of respondents indicated they would like to get a COVID-19 vaccine, while 74.3% of respondents were either hesitant or completely rejected vaccination. The vaccine acceptance increased with increasing age, education, and income level. Major motivation of pro-vaccination behaviour was intention to achieve collective immunity (30.1%), while the leading incentive for vaccine refusal was deficiency of clinical data (30.2%). The Pfizer-BioNTech vaccine is shown to be eightfold more preferred vaccine compared to the other manufacturers. For the first time, vaccine acceptance among health care professionals has been reported, where only 39.4% of healthcare professionals expressed willingness to get vaccinated.

With the high share of the population unwilling to vaccinate, governmental impotence in securing the vaccines supplies, combined with the lack of any lockdown measures suggests that Bosnia and Herzegovina is unlikely to put COVID-19 pandemic under control in near future.

## Introduction

On 1st March 2020, the World Health Organization (WHO) characterized the coronavirus disease 2019 (COVID-19) as a pandemic (https://www.who.int/director-general/speeches/detail/who-director-general-s-opening-remarks-at-the-media-briefing-on-covid-19---11-march-2020). Since the first registered case of COVID-19 until now there were more than 100 million officially registered cases of COVID-19 and more than 2 million persons have passed away due to COVID-19 infection (https://www.worldometers.info/coronavirus/). Consequently, the rapid development of a COVID-19 vaccine was a global imperative^1^. Now in 2021, there are currently a few vaccines that passed the third phase of clinical trial and they are being distributed all over the world^2^. In the majority of developed countries, the vaccination has already started, whereas in most developing and less developed countries the vaccination has not yet started (https://ourworldindata.org/covid-vaccinations).

Vaccination has already started in the United States (US) and the European Union (EU). In the US and EU, Pfizer-BioNTech and Moderna vaccines have been approved (https://www.pfizer.com/news/press-release/press-release-detail/pfizer-and-biontech-receive-authorization-european-union),while European Medicines Agency (EMA) has recommended the approval of the AstraZeneca COVID-19 vaccine (https://www.ema.europa.eu/en/news/ema-recommends-first-covid-19-vaccine-authorisation-eu). Safety and efficiency of the COVID-19 vaccine has been also confirmed for the Sputnik V.^3^ Additionally, the National Medical Products Administration in China has given approval for the COVID-19 vaccine made by Sinovac Biotech (http://subsites.chinadaily.com.cn/nmpa/2021-02/07/c_588422.htm). Regarding the Balkans, vaccination has not started in Albania, Bosnia and Herzegovina (B&H), Kosovo, Montenegro, and North Macedonia (Figure 1). In B&H, the media have reported that in Republika Srpska, an entity of Bosnia and Herzegovina, around 2.000 doses of Sputnik V COVID-19 vaccine have been distributed among healthcare workers.

**Figure 1.**
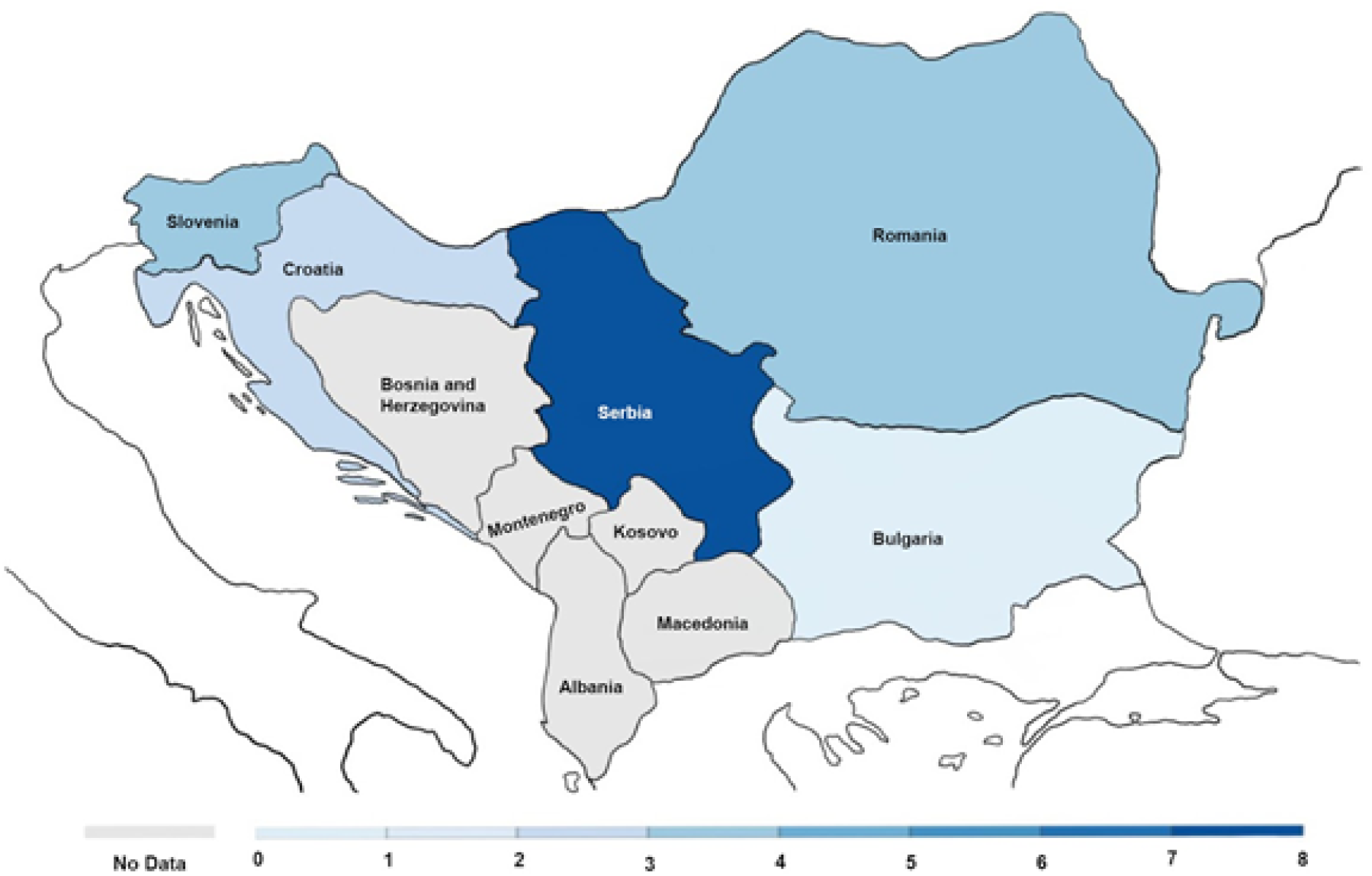
The number of vaccine doses given to people, not the number of people fully vaccinated. Since some vaccines require more than one dose, the number of fully vaccinated people is likely to be lower. Retrieved from Our World in Data on 2/3/2021.

In Bosnia and Herzegovina, ∼120.000 cases of COVID-19 have been officially been registered until February 2021 (3.4% of the whole population) and almost 5.000 deaths (4.16% of all COVID-19 cases). The peak of infection was in October and November. Currently, in B&H there are around 400 active cases, with ∼93 new confirmed cases daily per million people and ∼4 deaths daily per million people (https://ourworldindata.org/coronavirus-data-explorer). In January 2021, B&H was 4th among the countries with the highest mortality rate due to COVID-19 infection with 123 deaths reported per 100,000 people (https://worldmapper.org/maps/coronavirus-cases-casemortality/).

Even though some COVID-19 measures are present (the curfew from 23:00 to 5:00 h, the ban of public gatherings for >50 persons indoors and 100 persons outdoors), they are not enforced. There is no lock-down, borders are open, schools and universities are partially opened, while shopping centers, restaurants, ski centers, and bars are working as usual (https://www.dw.com/bs/njema%C4%8Dki-mediji-gra%C4%91ani-bih-se-ne-pridr%C5%BEavaju-pravila/a-54047892).

The aim of this study was to collect data about the willingness of the adult population in Bosnia and Herzegovina to be vaccinated and to examine the factors that affect vaccine rejection. Additionally, we examined if vaccine rejection was correlated with education, income, profession, and age.

## Materials and Methods

We conducted a cross-sectional electronic survey study about COVID-19 vaccine acceptance in Bosnia and Herzegovina from January 26^th^ to February 2^nd^, 2021 gathering answers from a total of 10,471 participants. The study was approved by the Ethics Committee of the Faculty of Engineering and Natural Sciences, International Burch University. Eligibility criteria included being age 18 or older and currently living in Bosnia and Herzegovina. The survey was developed in the local language and created using Google’s online survey platform. All the study participants were informed that the data would be used only for research purposes and not available to the public. According to Google’s privacy policy, all survey responses were anonymous and confidential. It was delivered to respondents via e-mails, research and employment-oriented online services (ResearchGate™ and LinkedIn™), and other social media platforms such as Facebook™, Skype™, and Viber™).

The participants responded to a total of 11 items. The first part of the survey covered demographic questions including gender, level of education, profession, age, and monthly income. Gender was categorized as male, female or other. The level of education was defined as elementary school, high school, undergraduate degree, and postgraduate degree (master or doctorate). The profession was classified into five categories including medical professionals, teachers, business sector, catering and service industry, and others. The age was categorized into four different groups: 18-30, 31-50, 51-64, and 65 years or older. Monthly income was defined as 250 EUR or less, 250-450 EUR, and 450 EUR or more.

The second part of the survey assessed a range of vaccine-related questions. Respondents were asked to claim whether they will choose to vaccinate or not, and to corroborate their choice with rationale for or against vaccination having the ability to select multiple options. Furthermore, participants were asked to state their major source of information about health implications of COVID-19 vaccines. The respondents willing to be vaccinated were asked to indicate which vaccine manufacturer(s) would be their personal choice: Pfizer – BioNTech (Germany), Oxford-AstraZeneca (United Kingdom), Modern (USA), Sputnik V (Russia), or Sinovac (China), and to choose one or more reasons for the choice.

Statistical analysis included descriptive statistics data regarding the frequencies calculated for each category of demographic set of questions. Also, a univariate and multinomial logistic regressions in R were employed to examine correlation between vaccine acceptability and a set of demographics and variables of interest.

## Results

Table 1 summarizes the set of demographic data including age, gender, education, monthly income, and profession. Women were 52.3% respondents of the survey and 53.9% were between 18 and 30 years old. More than half of the participants (53.1%) had monthly income of 450 EUR or more (average salary is about 450 EUR). About half of the respondents (51.9%) had a university degree. Significant number of healthcare professionals (15%) took part in our study.

**Table 1.** Summary of participants’ demographic data

|  |  |
| --- | --- |
|  | **Overall** |
| **n** | **10,471** |
| **Gender (%)** |  |
| Male | 4,965 (47.4) |
| Female | 5,476 (52.3) |
| Other | 30 (0.3) |
| **Level of education (%)** |  |
| Elementary school | 159 (1.5) |
| High school | 4,878 (46.6) |
| Undergraduate degree | 3,757 (35.9) |
| Postgraduate degree (Master or Doctoral degree) | 1,677 (16) |
| **Profession (%)** |  |
| Medical professionals | 1,570 (15) |
| Education sector | 936 (8.9) |
| Economic sector | 1,639 (15.7) |
| Catering and service industry | 721 (6.9) |
| Other | 5,605 (53.5) |
| **Age group in years (%)** |  |
| 18-30 | 5,649 (53.9) |
| 31-50 | 4,210 (40.2) |
| 51-64 | 544 (5.2) |
| 65+ | 68 (0.6) |
| **Total monthly income** |  |
| 250 EUR or less | 2,522 (24.1) |
| 250-450 EUR | 2,384 (22.8) |
| 450 EUR or more | 5,565 (53.1) |

Overall, 25.7% (2,695 of 10,461) of respondents indicated they are willing to get a COVID-19 vaccine, while 74.3% of respondents hesitated to get vaccinated (37.4% would not vaccinate, 13.7% respondents would vaccinate only if obliged, and 23.2% will wait for additional clinical studies to decide). Detailed breakdown of vaccine questions is available in Supplementary Table 1. We treated the three answers: ‘No’, ‘Only if I will have to’ and ‘Maybe later’ as one group because they show trends in their answers (Supplementary Table 2 and 3).

Table 2 summarizes 5 univariate regressions regarding vaccine acceptability against demographics (age, gender, monthly income, education, and profession). Accordingly, age, education, occupation and income significantly affected attitudes towards vaccination (p <.05), while sex of the participant did not (p > .05). People aged 31–50, 51–64 and 65+ were more likely to accept the vaccine than those who were aged 18–30. This difference was strongest (odds ratio (OR) = 4.61; 95% confidence interval (CI) (2.74, 7.77)) when respondents aged 65+ were compared to the youngest age cohort. The univariate regression suggests no significant distinction in the response to vaccine acceptance based on the gender.

**Table 2.**
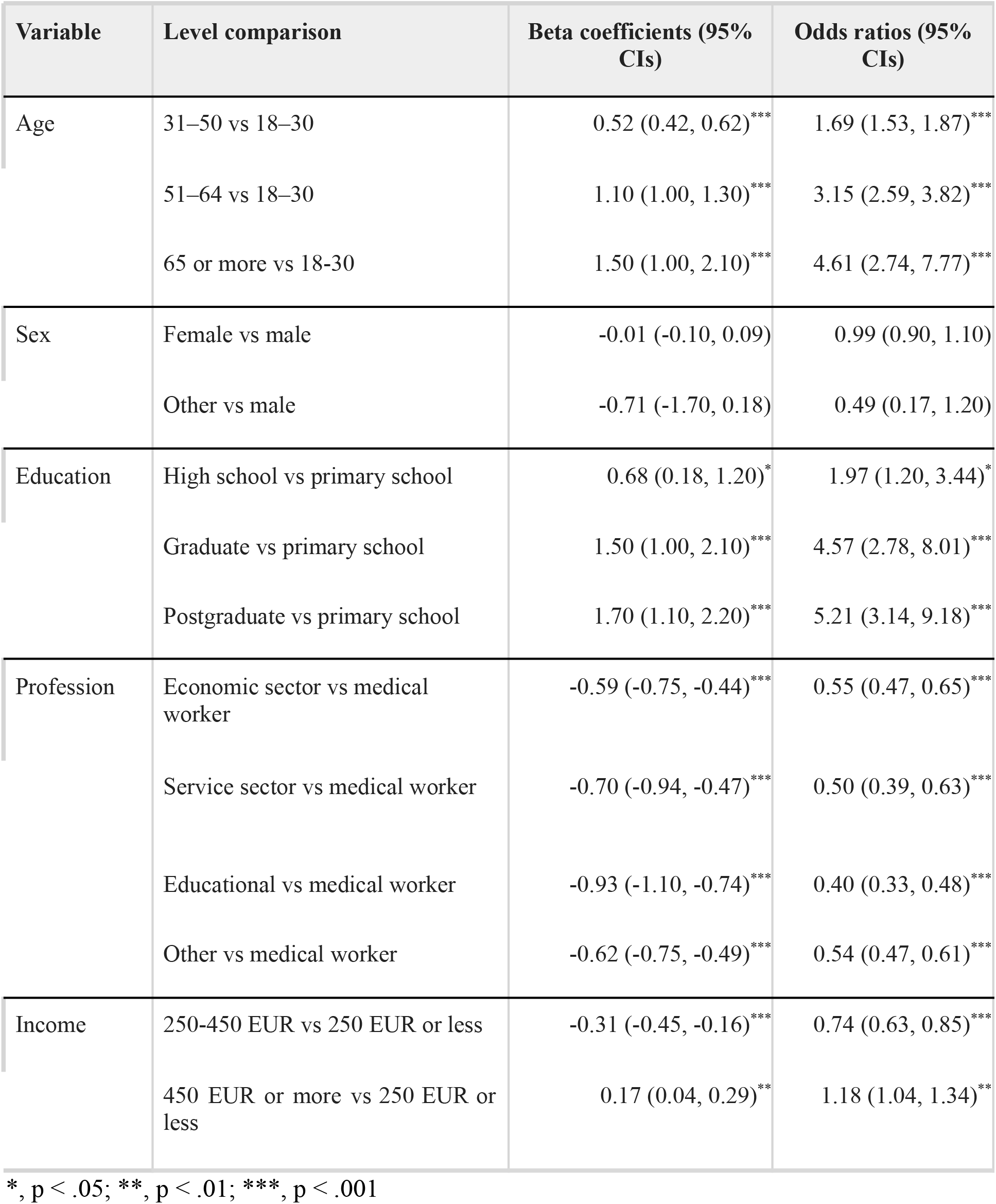
Beta coefficients and odds ratios of predictors of attitude towards vaccination when comparing the answers ‘Yes’ with ‘No / Only if I will have to / Maybe later’

| Variable | Level comparison | Beta coefficients (95% CIs) | Odds ratios (95% CIs) |
| --- | --- | --- | --- |
| Age | 31–50 vs 18–30 | 0.52 (0.42, 0.62)*** | 1.69 (1.53, 1.87)*** |
|  | 51–64 vs 18–30 | 1.10 (1.00, 1.30)*** | 3.15 (2.59, 3.82)*** |
|  | 65 or more vs 18–30 | 1.50 (1.00, 2.10)*** | 4.61 (2.74, 7.77)*** |
| Sex | Female vs male | -0.01 (-0.10, 0.09) | 0.99 (0.90, 1.10) |
|  | Other vs male | -0.71 (-1.70, 0.18) | 0.49 (0.17, 1.20) |
| Education | High school vs primary school | 0.68 (0.18, 1.20)* | 1.97 (1.20, 3.44)* |
|  | Graduate vs primary school | 1.50 (1.00, 2.10)*** | 4.57 (2.78, 8.01)*** |
|  | Postgraduate vs primary school | 1.70 (1.10, 2.20)*** | 5.21 (3.14, 9.18)*** |
| Profession | Economic sector vs medical worker | -0.59 (-0.75, -0.44)*** | 0.55 (0.47, 0.65)*** |
|  | Service sector vs medical worker | -0.70 (-0.94, -0.47)*** | 0.50 (0.39, 0.63)*** |
|  | Educational vs medical worker | -0.93 (-1.10, -0.74)*** | 0.40 (0.33, 0.48)*** |
|  | Other vs medical worker | -0.62 (-0.75, -0.49)*** | 0.54 (0.47, 0.61)*** |
| Income | 250-450 EUR vs 250 EUR or less | -0.31 (-0.45, -0.16)*** | 0.74 (0.63, 0.85)*** |
|  | 450 EUR or more vs 250 EUR or less | 0.17 (0.04, 0.29)** | 1.18 (1.04, 1.34)** |

\*, $p < .05$ ; \*\*, $p < .01$ ; \*\*\*, $p < .001$

Higher income was positively associated with vaccine acceptance. People earning 450+ EUR per month were 1.18 (95 CI% (1.04, 1.34)) times more likely to respond positively to the vaccine acceptance question than people earning 250 EUR and less. Higher levels of education were also associated positively with vaccine acceptance. Respondents from the postgraduate group were 5.21 (95 CI% (3.14, 9.18)) times more likely to respond positively compared to participants having only primary school education. Medical health professionals were more likely to get vaccinated compared to other professions. In fact, educational workers had 60% lower odds of vaccine acceptance compared to the health professionals.

Major determinants behind vaccination were achieving collective immunity (30.11%) and concern regarding personal health (29.57%), following avoidance of “travel ban” (27.31%) and employer request (13.00%). The Pfizer-BioNTech would be chosen by 50.62 % participants willing to vaccinate, while Sinovac vaccines would be preferred for only 6.44 % of them. Effectiveness shown in clinical trials is the main motive for Pfizer’s vaccine choice. Most objections to vaccination are due to insufficient clinical trials (30.11 %), 23.08 % respondents perceive pharmaceutical companies as self-serving enterprises. Significant numbers recognize vaccines as harmful (12.23%), 9.63% participants identify COVID-19 disease as harmless to their health, while an identical portion of respondents reject vaccines due to religious motives. For 9.19% participants SARS-CoV-2 virus is just a conspiracy theory, while 6.05% individuals assessed vaccines as necessary only for clinically vulnerable citizens.

## Discussion

In this study, we report the lowest COVID-19 vaccine acceptance in the world, where only 25.7% participants demonstrated willingness to receive vaccination against SARS-CoV-2. Lowest COVID-19 vaccination acceptance levels prior reported was in Poland (37 %), following Slovakia (41 %), Romania (44 %) and Czech Republic (49 %)^4-5^. Understanding of vaccination refusal and reasons for rejection among citizens in B&H is of great importance as reports from the January 2021 list Bosnia and Herzegovina as fourth in the world in terms of deaths per 100,000 inhabitants, right after Slovenia, Belgium and San Marino (https://worldmapper.org/maps/coronavirus-cases-casemortality/). Observed data should be used to raise awareness among the population and reach those strongly advocating against COVID-19 vaccination programs.

Univariate regression outputs for vaccine acceptability demonstrate important discrepancies across diverse categories in the survey. Participants with above average income were more likely to accept vaccination compared to those having minimum wage. Findings suggest participants with primary school education were more prone to reject vaccination compared to participants having higher levels of education. Observed data are in accordance with studies previously conducted^4^.

The univariate regression suggests no significant distinction in the response to vaccine acceptance based on the gender. However, we see a trend where women seem to be more hesitant regarding COVID-19 vaccines, while men are slightly more prone to vaccination, diverting from the trend of higher medical care service utilization among women^6^. Additionally, we observed age-related associations with vaccine acceptance. Older people were more likely to report that they would take a vaccine, whereas respondents aged 18−30 years had the highest rate of vaccination refusal^5,7,8^.

For the first time in B&H, vaccine acceptance among health care professionals has been examined and compared to the other professions. Only 39.4 % of healthcare professionals are willing to accept vaccination, while others are hesitant or strongly refusing vaccination. This confirms concerns raised by Arapovic et al. in 2019, regarding lower vaccine acceptance among healthcare workers in B&H, as they directly communicate with patients and shape their perspective toward vaccination^9^. Similar vaccine acceptance was reported in a recent study conducted among health care professionals in the United States^10^.

Major driver of pro-vaccination behaviour was intention to achieve collective immunity, following health care and personal protection. Also, data clearly shows employer’s vaccination requests would be insufficient incentive for vaccine acceptance among employees. The participants willing to vaccinate prefer Pfizer-BioNTech vaccines up to eightfold more compared to the other vaccine manufacturers, acknowledging high vaccine effectiveness reported in clinical trials^11-15^. Confidence in system and governmental decisions is evidently low, as the population witnesses various political and socio-economic crises in the post-war period. Strong domestic anti-vaccination movement noticed in the last several years finally got better understanding through cross-examination and common objections anti-vaccine advocates expressed over the years are reported in our survey^16,17^. Anti-vaccination groups target local media and online platforms to spread misleading health information and address controversial arguments such as the economic benefit of pharmacies and tragic personal stories^18^. Reporting educational programs, media platforms and social networks as main sources of information during pandemic, high COVID-19 vaccine rejection among participants becomes utterly understandable. As the second major motivation for vaccine rejection, participants listed mistrust in pharmaceutical companies, followed by assessment of vaccines as harmful. Scientific community and health care professionals advocating vaccines, must be more presented on those platforms to raise awareness and reach citizens looking for reliable information.

Most of the data used in this survey have been collected using online social networks, which often excludes citizens in the category of age 65 and older^19^. Since they represent a high-risk group and are more likely to accept vaccination, the acceptance rate may be larger than presented^4,5^. Another limitation represents absence of information in case participants were infected with SARS-CoV-2 virus and whether they consider acquired immunity to be sufficient protection and adequate replacement for the vaccination. 23.2 % participants indicated hesitance to the vaccination due to insufficient clinical trials conducted, therefore safe and effective mass immunization around the globe could increase acceptance rate as time passes. Finally, rejection was assessed using a hypothetical vaccine, which may differ from the respondents’ preferences encountering real life situations once COVID 19 vaccines become widely available.

According to current studies, herd immunity benefits are achievable if 65%–70% of the population is vaccinated^20^. With the high share of the population unwilling to vaccinate, governmental impotence in securing the vaccines’ supplies, combined with the number of people unable to receive the COVID-19 vaccine (e.g., allergies), herd immunity is out of reach for the B&H population in the near future. In order to increase awareness regarding health benefits of vaccination and the historical role immunization had in eradication of many deadly diseases, people must be reached through main informing sources - educational programs and media. Additional efforts must be made to organize scientific panels and conferences for healthcare workers and physicians, as only 39,4% of them are willing to accept vaccination. Ideally, frontline medical professionals should make strong recommendations for vaccination, as well as share their personal experiences with COVID-19 vaccines. Finally, preparation for public acceptance of a COVID-19 vaccine must be carefully conducted before a vaccine becomes widely available. Based on this study, we urge the Bosnian government to develop strategies and COVID-19 vaccination implementation plans that would encourage citizens to accept a vaccination^21^.

## Data Availability

Data available within the article or its supplementary materials.

## Contributions

A.F. and A.O. conceived and designed the study. N.Đ. and A.F. managed and performed data collection. E.M. and I.M. statistically analyzed and interpreted the data. A.F., A.O., N. Đ., A.F., Z.G., and A.K. drafted the manuscript. A.K. edited and approved the final version for submission.

## Ethical approval

This study was approved by Burch University Ethics Commission and informed consent obtained from all participants.

---

**Supplementary Table 1.** Summary of descriptive statistics results

| Research Questions | n=10,471 |
| --- | --- |
| Will most of the people reject the COVID-19 vaccine? | 37.40% does not want to receive COVID-19 vaccine<br>23.18% will be waiting for additional clinic studies<br>13.69% will receive only if obliged to |
| What are the main motives for vaccination? | 30.11% Acquiring collective immunity and preventing spread of virus<br>29.57% Healthcare and self protection<br>27.31% Travel possibility and avoidance of „travel ban“<br>13.00% Employer’s request and preservation of working position |
| What are the reasons for vaccination refusal? | 30.19% Insufficient clinical trials of vaccination<br>23.08% Lack of trust to pharmacy<br>12.23% Vaccines perceived as harmful<br>9.63% COVID-19 disease is not dangerous for my health<br>9.63% Vaccines’ compositions are against my ethical and religious principles<br>9.19% SARS-CoV-2 does not exist - it is conspiracy theory<br>6.05% Only most vulnerable categories shall receive the vaccines |
| What are the main sources of information about health-related implication vaccines have? | 27.70% Educational and documentary shows<br>26.16% Media and information portals<br>20.10% Social networks (Facebook™, Twitter™ etc.)<br>15.90% Scientific books and papers<br>10.14% Recommendations of family physicians |
| Which vaccines are more likely to be accepted by citizens of Bosnia and Herzegovina? | No1: Pfizer–BioNTech (Germany), by 50.62%<br>No2: Oxford-AstraZeneca (United Kingdom), by 16.16%<br>No3: Moderna (USA), by 15.30%<br>No4: Sputnik V (Russia), 11.48%<br>No5: Sinovac (China), by 6.44% |
| What are the reasons to choose a specific vaccine manufacturer? | 52.37% Effectiveness reported through studies<br>21.69% International politics states' manufacturers have<br>19.92% Physician's or medical staff's advice<br>6% Following governmental decisions |

**Supplementary Table 2.** Beta coefficients and odds ratios of predictors of attitude towards vaccination when comparing the vaccine answers using multinomial logistic regression

| Characteristic | Beta | 95% CI | Odds Ratio | 95% CI | p-value |
| --- | --- | --- | --- | --- | --- |
| **Yes vs. No** |  |  |  |  |  |
| Female vs. Male | 0.10 | -0.01, 0.21 | 1.11 | 0.99, 1.23 | 0.074 |
| Other vs. Male | -1.10 | -2.10, -0.14 | 0.32 | 0.12, 0.87 | 0.025 |
| ----- |  |  |  |  |  |
| Postgraduate vs. primary school | 2.20 | 1.60, 2.70 | 8.72 | 5.01, 15.2 | <0.001 |
| Graduate vs. primary school | 1.90 | 1.40, 2.50 | 6.99 | 4.06, 12.0 | <0.001 |
| High school vs. primary school | 0.87 | 0.33, 1.4 | 2.38 | 1.38, 4.08 | 0.002 |
| ----- |  |  |  |  |  |
| Economic sector vs. medical worker | -0.63 | -0.81, -0.44 | 0.53 | 0.44, 0.64 | <0.001 |
| Other vs. medical worker | -0.74 | -0.89, -0.59 | 0.48 | 0.41, 0.55 | <0.001 |
| Service sector vs. medical worker | -0.81 | -1.10, -0.55 | 0.45 | 0.35, 0.58 | <0.001 |
| Educational worker vs. medical worker | -1.00 | -1.20, -0.73 | 0.39 | 0.31, 0.48 | <0.001 |
| ----- |  |  |  |  |  |
| 65 or more vs 18-30 | 1.50 | 0.85, 2.1 | 4.27 | 2.33, 7.81 | <0.001 |
| 51-64 vs 18-30 | 1.30 | 1.10, 1.60 | 3.81 | 3.01, 4.82 | <0.001 |
| 31-50 vs 18-30 | 0.59 | 0.48, 0.70 | 1.81 | 1.62, 2.02 | <0.001 |
| ----- |  |  |  |  |  |
| 450 EUR or more vs 250 EUR or less | 0.09 | -0.05, 0.23 | 1.10 | 0.95, 1.26 | 0.200 |
| 250 EUR - 450 EUR vs 250 EUR or less | -0.43 | -0.59, -0.27 | 0.65 | 0.55, 0.76 | <0.001 |
| **Only if having to vs. No** |  |  |  |  |  |
| Female vs. Male | -0.04 | -0.17, 0.09 | 0.96 | 0.85, 1.10 | 0.600 |
| Other vs. Male | -11.00 | -11.00, -11.00 | 0.00 | 0.00, 0.00 | <0.001 |
| Postgraduate vs. primary school | 0.39 | -0.12, 0.90 | 1.48 | 0.89, 2.47 | 0.130 |
| Graduate vs. primary school | 0.45 | -0.03, 0.94 | 1.57 | 0.97, 2.56 | 0.068 |
| High school vs. primary school | 0.15 | -0.33, 0.63 | 1.16 | 0.72, 1.87 | 0.600 |
| Economic sector vs. medical worker | 0.00 | -0.23, 0.24 | 1.00 | 0.79, 1.27 | >0.900 |
| Other vs. medical worker | -0.24 | -0.44, -0.04 | 0.79 | 0.65, 0.96 | 0.018 |
| Service sector vs. medical worker | -0.07 | -0.35, 0.21 | 0.93 | 0.70, 1.23 | 0.600 |
| Educational worker vs. medical worker | 0.31 | 0.05, 0.58 | 1.37 | 1.05, 1.78 | 0.020 |
| 65 or more vs 18-30 | -0.85 | -2.10, 0.38 | 0.43 | 0.13, 1.46 | 0.200 |
| 51-64 vs 18-30 | -0.06 | -0.41, 0.28 | 0.94 | 0.67, 1.32 | 0.700 |
| 31-50 vs 18-30 | 0.04 | -0.09, 0.17 | 1.04 | 0.91, 1.18 | 0.600 |
| 450 EUR or more vs 250 EUR or less | 0.05 | -0.11, 0.22 | 1.06 | 0.90, 1.24 | 0.500 |
| 250 EUR - 450 EUR vs 250 EUR or less | 0.01 | -0.17, 0.19 | 1.01 | 0.85, 1.20 | >0.900 |
| **Maybe later vs. No** |  |  |  |  |  |
| Female vs. Male | 0.33 | 0.22, 0.44 | 1.39 | 1.25, 1.56 | <0.001 |
| Other vs. Male | -0.64 | -1.6, 0.31 | 0.53 | 0.20, 1.36 | 0.200 |
| Postgraduate vs. primary school | 1.50 | 1.00, 1.90 | 4.39 | 2.75, 7.02 | <0.001 |
| Graduate vs. primary school | 1.20 | 0.77, 1.7 | 3.39 | 2.15, 5.34 | <0.001 |
| High school vs. primary school | 0.66 | 0.21, 1.1 | 1.94 | 1.23, 3.03 | 0.004 |
| Economic sector vs. medical worker | -0.08 | -0.28, 0.11 | 0.92 | 0.75, 1.12 | 0.400 |
| Other vs. medical worker | -0.22 | -0.38, -0.05 | 0.81 | 0.69, 0.95 | 0.009 |
| Service sector vs. medical worker | -0.27 | -0.51, -0.02 | 0.77 | 0.60, 0.98 | 0.035 |
| Educational worker vs. medical worker | -0.26 | -0.49, -0.03 | 0.77 | 0.61, 0.97 | 0.028 |
| 65 or more vs 18-30 | 0.10 | -0.65, 0.85 | 1.10 | 0.52, 2.34 | 0.800 |
| 51-64 vs 18-30 | 0.56 | 0.31, 0.81 | 1.75 | 1.36, 2.25 | <0.001 |
| 31-50 vs 18-30 | 0.19 | 0.07, 0.30 | 1.20 | 1.08, 1.35 | 0.001 |
| 450 EUR or more vs 250 EUR or less | -0.25 | -0.39, -0.12 | 0.78 | 0.68, 0.89 | <0.001 |
| 250 EUR - 450 EUR vs 250 EUR or less | -0.38 | -0.53, -0.24 | 0.68 | 0.59, 0.79 | <0.001 |

**Supplementary Table 3.** Beta coefficients and odds ratios of predictors of attitude towards vaccination when comparing the answers ‘Yes / Only if I will have to’ <u>vs.</u> ‘No / Maybe later’

| Variable | Level comparison | Beta coefficients (95% CIs) | Odds ratios (95% CIs) |
| --- | --- | --- | --- |
| Age | 31–50 vs 18–30 | 0.31 (0.23, 0.40)*** | 1.37 (1.25, 1.49)*** |
|  | 51–64 vs 18–30 | 0.66 (0.48, 0.85)*** | 1.94 (1.62, 2.33)*** |
|  | 65 or more vs 18-30 | 0.84 (0.34, 1.30)** | 2.31 (1.40, 3.84)*** |
| Sex | Female vs male | -0.08 (-0.17, 0.00) | 0.92 (0.84, 1.00) |
|  | Other vs male | -1.2 (-2.2, -0.35)* | 0.30 (0.11, 0.71)* |
| Education | High school vs primary school | 0.33 (-0.03, 0.72) | 1.39 (0.97, 2.05) |
|  | Graduate vs primary school | 0.91 (0.54, 1.30)*** | 2.48 (1.72, 3.66)*** |
|  | Postgraduate vs primary school | 0.94 (0.56, 1.30)*** | 2.56 (1.75, 3.80)*** |
| Profession | Economic sector vs medical worker | -0.40 (-0.55, -0.26)*** | 0.67 (0.58, 0.77)*** |
|  | Service sector vs medical worker | -0.44 (-0.64, -0.25)*** | 0.64 (0.53, 0.78)*** |
|  | Educational vs medical worker | -0.42 (-0.59, -0.26)*** | 0.65 (0.55, 0.77)*** |
|  | Other vs medical worker | -0.51 (-0.62, -0.39)*** | 0.60 (0.54, 0.68)*** |
| Income | 250-450 EUR vs 250 EUR or less | -0.09 (-0.21, 0.04) | 0.92 (0.81, 1.04) |
|  | 450 EUR or more vs 250 EUR or less | 0.19 (0.08, 0.30)*** | 1.21 (1.09, 1.35)*** |

\*, $p < .05$ ; \*\*, $p < .01$ ; \*\*\*, $p < .001$

